# An interactive COVID-19 virus Mutation Tracker (CovMT) with a particular focus on critical mutations in the Receptor Binding Domain (RBD) region of the Spike protein

**DOI:** 10.1101/2021.01.22.21249716

**Authors:** Intikhab Alam, Aleksandar Radovanovic, Roberto Incitti, Allan Kamau, Muhammad Alarawi, Esam I. Azhar, Takashi Gojobori

## Abstract

Almost one year has passed since the appearance of SARS-CoV-2, causing the COVID-19 pandemic. The number of confirmed SARS-Cov-2 cases worldwide has now reached ∼92 million, with 2 million reported deaths (https://covid19.who.int). Nearly 400,000 SARS-Cov-2 genomes were sequenced from COVID-19 samples and added to public resources such as GISAID (https://gisaid.org). With the vaccines becoming available or entering trials (https://covid19.trackvaccines.org), it is vital to keep track of mutations in the genome of SARS-CoV-2, especially in the Spike protein’s Receptor Binding Domain (RBD) region, which could have a potential impact on disease severity and treatment strategies.^1–3^ In the wake of a recent increase in cases with a potentially more infective RBD mutation (N501Y) in the United Kingdom, countries worldwide are concerned about the spread of this or similar variants. Impressive sampling and timely increase in sequencing efforts related to COVID-19 in the United Kingdom (UK) helped detect and monitor the spread of the new N501Y variant. Similar sequencing efforts are needed in other countries for timely tracking of this or different variants. To track geographic sequencing efforts and mutations, with a particular focus on RBD region of the Spike protein, we present our daily updated COVID-19 virus Mutation Tracker system, see https://www.cbrc.kaust.edu.sa/covmt.

## Main

We developed a daily updated COVID-19 virus Mutation Tracker system (CovMT, see https://www.cbrc.kaust.edu.sa/covmt and Figure 1A) on SARS-Cov-2 isolate genomes deposited to GISAID resource (http://gisaid.org), to track the worldwide sequencing efforts and the evolution of the mutational landscape of this virus. CovMT summarizes mutations from nearly 400,000 isolates into groups of generic virus clades, lineages, and more specific mutation sets we call Mutation Fingerprints (MF). These summaries, including associated metadata of location, date of sampling, and patient disease severity information, where available, at the continent and country levels, are accessible from the main page of the CovMT system.

**Figure 1:**
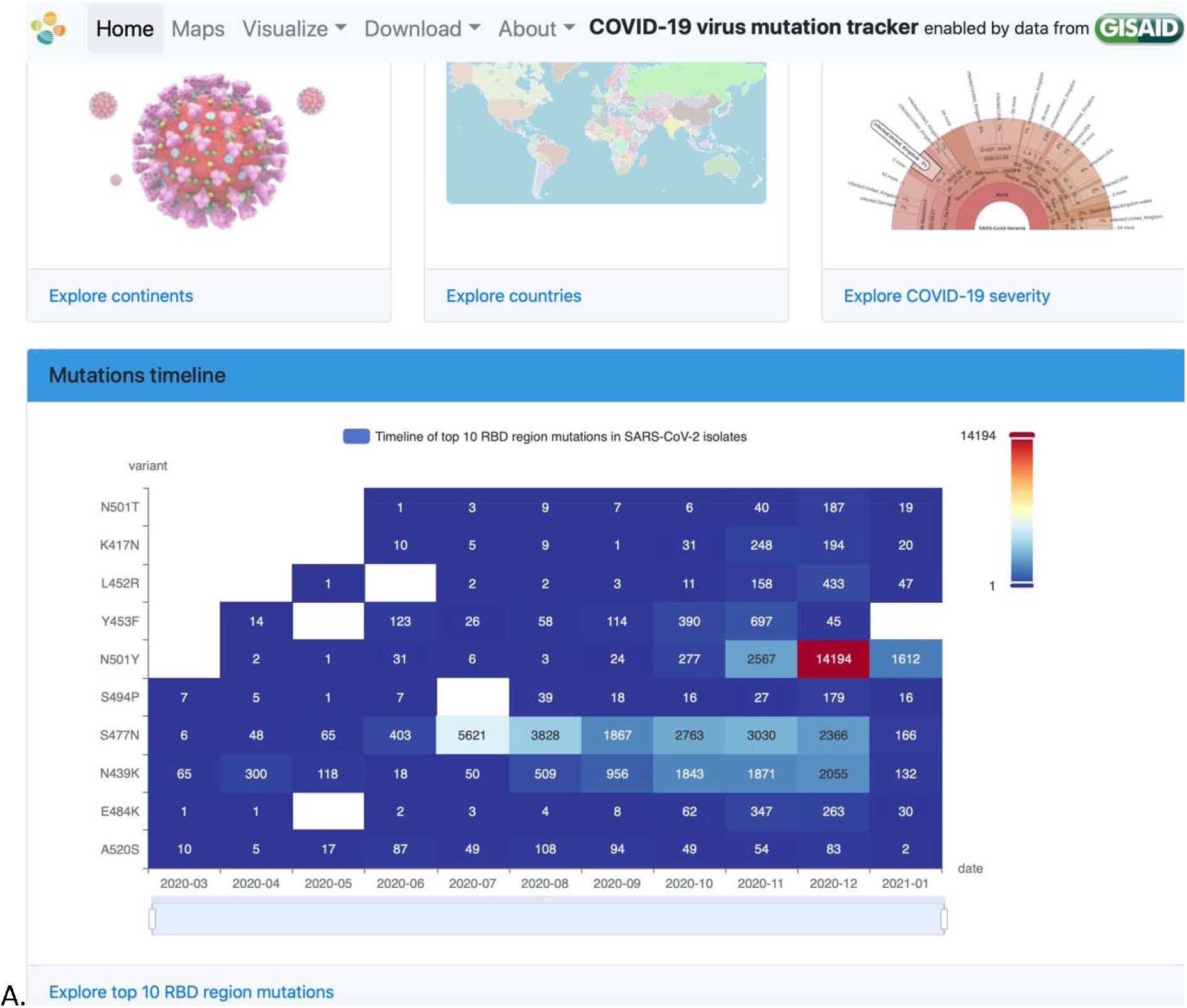
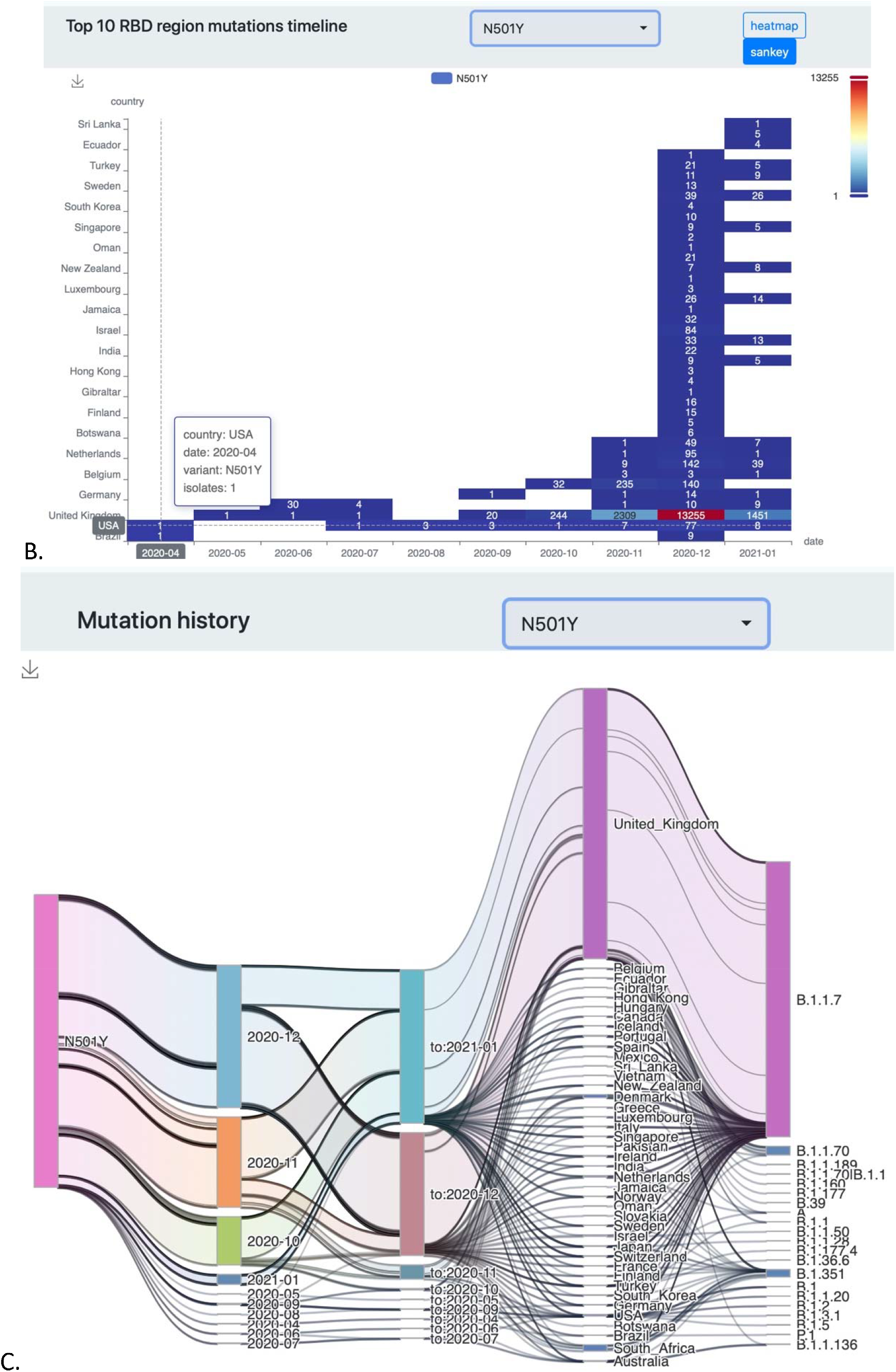
CovMT overview A) the Main page of CovMT showing options to explore mutations spread geographically (by continent and country), with related disease severity information and a timeline of the top mutations in the Spike protein RBD region. B) Timeline of N501Y mutation appearing in SARS-CoV-2 isolates in different countries. C. History of N501Y mutation with months (first seen, last seen), country and related lineages.

CovMT also provides a particular focus on the timeline of the spread of SARS-CoV-2 variants related to mutations in the RBD region of the Spike protein. Figure 1B shows a timeline graph of how the N501Y mutation appeared for the first time in April in Brazil and the USA. It also shows the number of isolate genomes significantly increasing after September 2020 when the N501Y UK variant, B.1.1.7, was detected. As of January 2020, the spread of N501Y variants can be detected in SARS-CoV-2 isolate genomes from 40 additional countries, using CovMT. Nonsynonymous mutations in the RBD region have a high potential to be linked to increased binding efficiency, increased infectivity, and the potential to evade antibodies^1–3^. To track all such variants, mutations in the RBD region are ranked based on their appearance in the number of SARS-CoV-2 isolate genomes in the CovMT system. The CovMT timeline (Figure 1A) shows that N501Y, S477N, N439K, and Y453F mutations can now be detected in more than 25,600, 20,900, 8,500, and 1,400 isolates, respectively, worldwide. An important RBD mutation, E484K, likely to evade existing antibodies^4^, originally appeared in the Denmark during March 2020, is now on the rise in South Africa^4^ since October 2020. More than 300 isolates show triple mutations (K417N, E484K and N501Y, lineage B.1.351) in South Africa, with some isolates now detected in the UK and 14 other countries. Timelines for top 10 RBD mutations can be explored at CovMT, https://www.cbrc.kaust.edu.sa/covmt/index.php?p=top-rbd-variants and history of mutations lineages at https://www.cbrc.kaust.edu.sa/covmt/index.php?p=mutation-history.

We defined Mutation Fingerprints (MFs) as the set of mutations present in the general population with a frequency above 0.005%. This information provides a more specific representation of all synonymous and nonsynonymous mutations found in an isolate of SARS-CoV-2 to facilitate more specific geographic tracking of variants and project COVID-19 severity information.

Location, date of sampling, and patient disease severity information, where available, are summarized for an MF related to a set of isolates, see supplementary information. The geographic origin status of an MF is considered foreign for a country if the earliest found location linked to its MF is not in that country. We categorize patient disease severity status, labeled *evidence*.*host*, as either asymptomatic, symptomatic mild, symptomatic severe, deadly, or unknown. When there are isolates represented by the same MF but without *evidence*.*host*, we impute the disease severity status as evidence.MF. In this way, we are able to project evidence.host based patient disease severity status from available ∼7% to ∼20% of isolates worldwide (see global <u>COVID-19 severity information</u> as an interactive circular graph or <u>an interactive MF table with timeline graphs)</u>. Taking the example of South African triple mutant, B.1.351, around 319 unique MFs are available. Here, patient disease severity information is available for at least 11 MFs, shown as symptomatic mild, one of such MFs appeared in the UK, see supplementary.

In summary, CovMT provides daily updated tracking and timelines of geographic sequencing efforts, mutations organized into known clades, newly defined MFs linked to local or foreign variants, and disease severity information through an easy-to-use CovMT system. With a particular focus on critical mutations in the RBD region of spike protein and with an option to seamlessly accrue the clinical metadata, including disease severity, we believe that CovMT will be useful for scientists, general public, and authorities to explore country-specific information.

## Data Availability

All SARS-CoV-2 isolate sequence data are obtained from GISAID (gisaid.org) and our daily updated mutation tracking analysis is available at CovMT website: https://www.cbrc.kaust.edu.sa/covmt

## Declaration of interests

None declared.

## Acknowledgment

We are thankful to KAUST Information Technology (IT) and KAUST supercomputing laboratory (KSL) teams for maintaining the computational resources and help in publishing the CovMT website. We pay special thanks to GISAID for providing daily updates on sequenced isolates, worldwide. This work is supported by King Abdulaziz City of Science and Technology (KACST) grant for COVID-19 research, number 0004-002-01-20-5.

## Supplementary Materials

Here we explain an example Mutation Fingerprint (MF) taking the example of the South African triple RBD mutation variant (K417N, E484K and N501Y) now detected in the UK, as highlighted in the Figure S1 A, with an associated timeline of isolates shown in Figure S1 B, and spread to other countries (lineage B.1.351 highlighted) in Figure S1 C, below. The MFs are more specific as they are able to capture additional mutations not yet defined in known clades or lineages. This allows to group all SARS-CoV-2 isolates with exactly the same set of mutations in to one MF. Such an MF can then be exploited to track associated variants. Furthermore, an MF allows to assign disease severity information if one of the isolates in a MF group are available with original disease severity information, we call evidence.host.

**Figure S1:**
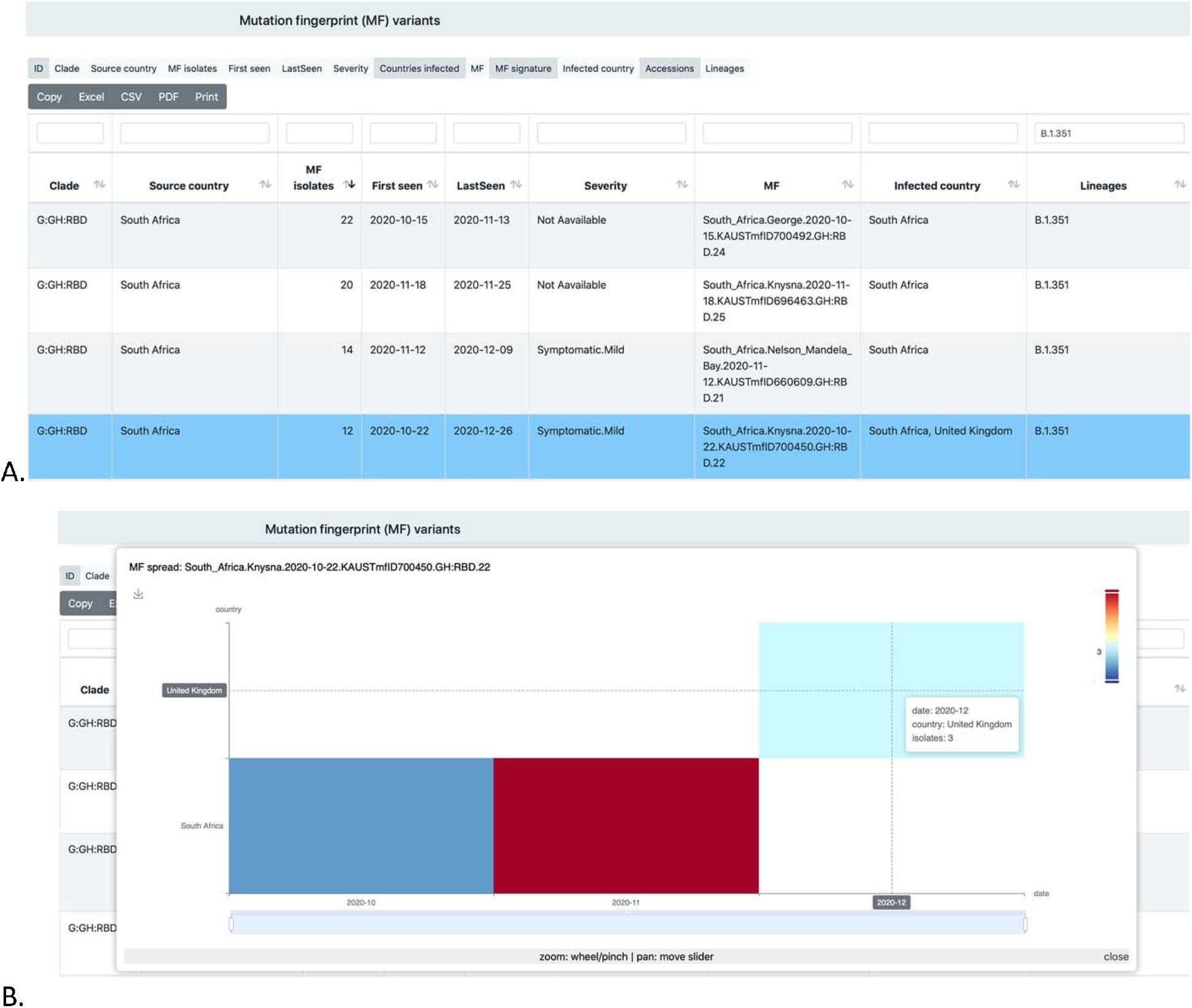
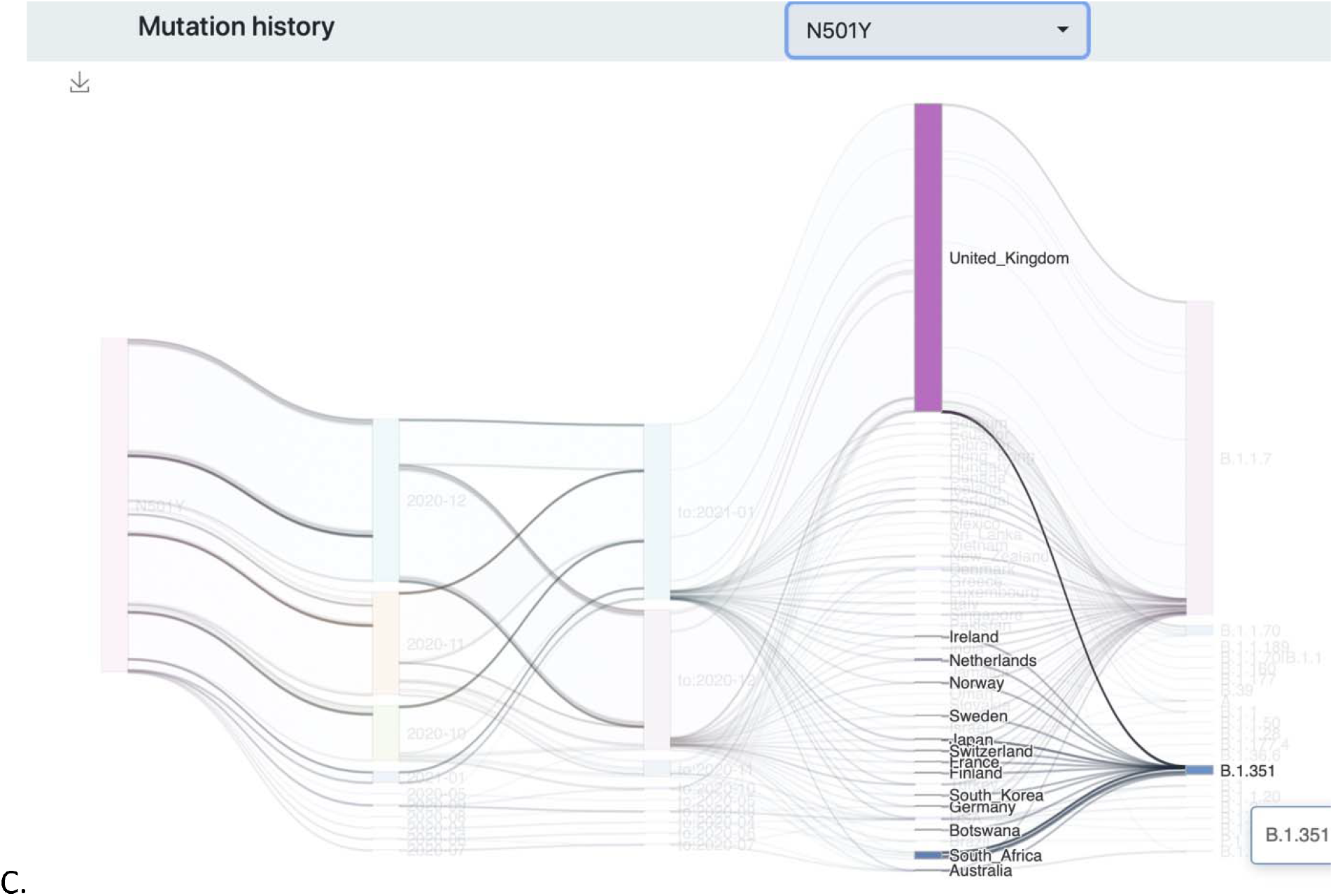
Mutation Fingerprint (MF) example of South African triple variant, lineage B.1.351. A) An interactive MF table showing SARS-CoV-2 clade, source country, first and last seen dates of an MF, disease severity information, MF ID, MF signature, and infected countries. B). Timeline of spread of South African variant, B.1.351, that appeared in the UK. Clicking a relevant MF table row shows a timeline graph for spread in related countries. C). History of B.1.351 with spread to several countries is shown.

A MF ID related to a single or set of isolates summarizes the location and date of earliest isolate, a unique internal id, followed by associated clade and number of mutations detected, as shown below:

Example MF ID: South_Africa.Knysna.2020-10-22.KAUSTmfID700450.GH:RBD.22

Mutations associated with an MF are recorded in a specific format, we call MF Signature, where all detected mutations for each gene are kept together, separated by a delimiter, “|”. For each gene, an MF signature shows the name of the gene or INTGEN for intergenic mutations, followed by the count of mutations and coordinates at the protein level as well as genome level, as shown below for the MF related to above shown MF ID:

E:1(P71L;C26456T)|INTGEN:2(INTGEN;G174T|INTGEN;C241T)|M:1(N41;C26645T)|N:1(T205I ;C28887T)|ORF3a:2(Q57H;G25563T|S171L;C25904T)|ORF8:1(F120;C28253T)|S:5(L18F;C216 14T|D80A;A21801C|D215G;A22206G|D614G;A23403G|A701V;C23664T)|S_RBD:3(K417N;G2 2813T|E484K;G23012A|N501Y;A23063T)|orf1ab:6(T265I;C1059T|T809;A2692T|F924;C3037T| K1655N;G5230T|K3353R;A10323G|P4715L;C14408T)

If we remove the coordinates of the mutations, a MF would show the count of mutations per gene, as shown below:

E:1|INTGEN:2|M:1|N:1|ORF3a:2|ORF8:1|S:5|S_RBD:3|orf1ab:6

All MFs are available for exploration at CovMT webpage: https://www.cbrc.kaust.edu.sa/covmt/index.php?p=world-variants

All MFs are available for exploration at CovMT mutant variant table (1). As an example, to search for the triple RBD mutant, one can use the text mentioned above, highlighted in yellow, and search the MF signature field in MF table (1).

We searched the mutation N501Y as a query in the MF signature field of MF table (1), and produced the following figure to show connections among first seen date, last seen date, country of isolate sampling and lineage information.

1. https://www.cbrc.kaust.edu.sa/covmt/index.php?p=world-variants

## Notes

### Competing Interest Statement

The authors have declared no competing interest.

